# Neoantigen DNA vaccines are safe, feasible, and capable of inducing neoantigen-specific immune responses in patients with triple negative breast cancer

**DOI:** 10.1101/2021.11.19.21266466

**Authors:** Xiuli Zhang, S. Peter Goedegebuure, Nancy B. Myers, Tammi Vickery, Michael D. McLellan, Feng Gao, Mark A. Sturmoski, Michael Y. Chen, Samuel W. Kim, Ina Chen, Jesse T Davidson, Narendra V. Sankpal, Jasreet Hundal, Lijin Li, Stephanie Myles, Rama Suresh, Cynthia X. Ma, Ademuyiwa Foluso, Andrea Wang-Gillam, Sherri Davies, Ian Hagemann, Elaine R. Mardis, Malachi Griffith, Christopher A. Miller, Ted H. Hansen, Timothy P. Fleming, Robert D. Schreiber, William E. Gillanders

## Abstract

**PURPOSE:** Cancer neoantigens are important targets of cancer immunotherapy. Neoantigen vaccines have the potential to induce or enhance highly specific antitumor immune responses with minimal risk of autoimmunity. We have developed a neoantigen DNA vaccine platform capable of efficiently presenting both HLA class I and II epitopes. To test the safety, feasibility and efficacy of this platform, we performed a phase 1 clinical trial in triple negative breast cancer patients with persistent disease following neoadjuvant chemotherapy, a patient population at high risk of disease recurrence.

**EXPERIMENTAL DESIGN:** Expressed somatic mutations were identified by tumor/normal exome sequencing and tumor RNA sequencing. The pVACtools software suite was used to identify and prioritize cancer neoantigens. Neoantigen DNA vaccines were designed and manufactured in an academic GMP facility at Washington University School of Medicine. Neoantigen DNA vaccines were administered via electroporation following completion of standard of care therapy. Safety was measured by clinical and laboratory evaluation. Immune responses were assessed by ELISPOT, flow cytometry and TCR sequencing.

**RESULTS:** 18 subjects received three doses of a personalized neoantigen DNA vaccine encoding on average 11 neoantigens per patient (range 4-20). The vaccinations were well tolerated with limited adverse events, primarily related to injection site reactions. Neoantigen-specific immune responses were induced in 16/18 patients as measured by ELISPOT and flow cytometry. At a median follow-up of 36 months, progression-free survival was 87.5% (95% CI: 72.7-100%) in the cohort of vaccinated patients compared to 49% (95% CI: 36.4-65.9%) in a cohort of institutional historical control patients (p=0.011).

**CONCLUSIONS:** Neoantigen DNA vaccines are safe, feasible, and capable of inducing a neoantigen-specific immune response. There is preliminary evidence of improved disease-free survival compared to historical controls.

## INTRODUCTION

Cancer neoantigens are mutant proteins/amino acid sequences expressed in tumors that can be recognized by the immune system. Cancer sequencing and related bioinformatics technologies have revolutionized our ability to identify cancer neoantigens. We performed one of the first preclinical studies to apply an immunogenomics approach to neoantigen identification [1]. This study demonstrated that cancer neoantigens are important targets of cancer immunoediting, and established the initial proof of concept that cancer exome sequencing and epitope prediction algorithms can be used to identify cancer neoantigens. In subsequent preclinical studies, we demonstrated that neoantigen vaccines can induce neoantigen-specific CD8 and CD4 T cell responses and antitumor immunity [2–5]. Other investigators have used similar strategies in the B16F10 (melanoma), TRAMP-C1 (prostate cancer), CT26, and MC-38 (colon cancer) mouse tumor models [6–8]. Because of the high mutational load and documented immunogenicity of human melanoma, initial clinical studies were carried out with melanoma patients using different vaccine platforms [9–11]. The first report of a neoantigen vaccine strategy in humans demonstrated that neoantigen dendritic cell vaccines are capable of generating neoantigen-specific T cell responses in human melanoma patients [9]. Two papers co-published in *Nature* by Ott *et al.* [11] and Sahin *et al.* [10] confirmed the potential of neoantigen vaccines in treating melanoma patients using neoantigen synthetic long peptide and RNA neoantigen vaccine approaches, respectively. More recent studies have evaluated neoantigen vaccines in glioblastoma [12, 13].

Triple-negative breast cancer (TNBC) lacks expression of estrogen receptor, progesterone receptor and HER2 gene amplification. TNBC is associated with an aggressive clinical course, and there are no targeted therapies available [14]. There is strong rationale to target cancer neoantigens in TNBC. First, TNBC is a mutationally complex breast cancer subtype [15–17]. The relative abundance of somatic mutations in TNBC suggests that neoantigens that can be targeted by neoantigen vaccine therapy are more likely to be present [17]. Second, tumor infiltrating lymphocytes (TILs) are more common in TNBC than in other breast cancer subtypes, and TILs are associated with improved outcome in TNBC following adjuvant, or neoadjuvant chemotherapy [18–20]. The association between TILs and improved outcome in TNBC highlights the importance of the adaptive immune system in the response to therapy. Third, several recent studies of chemotherapy combined with immune checkpoint inhibition in TNBC suggest that a percentage of patients with TNBC will benefit from combination immunotherapy with durable responses noted [21–24]. Although these studies are promising, there is clear room for improvement.

Our efforts towards improving clinical outcomes in TNBC have focused on a neoantigen DNA vaccine strategy. The observation that direct administration of recombinant DNA can generate potent immune responses established the field of DNA vaccines in the early 1990s [25–30]. Since that time, DNA vaccines have remained an area of intense research interest, and vaccines targeting infectious disease agents and cancers have progressed into clinical trials. The DNA vaccine platform affords flexibility by allowing targeting of multiple neoantigens using a single polyepitope DNA vaccine. We have designed such a neoantigen DNA vaccine platform that also integrates a mutant ubiquitin molecule in order to promote epitope generation and display [5]. In the present study, we used a TriGrid electroporation device to administer the neoantigen DNA vaccines. Electroporation dramatically increases DNA uptake by muscle cells, antigen expression, and immunogenicity [31–34]. Of particular note, electroporation has now been used successfully in non-human primates and in human clinical trials, with responses at levels not previously observed with other DNA vaccine approaches and similar to or superior to those induced by live vectors [35–45].

We have completed a phase 1 clinical trial of a neoantigen DNA vaccine strategy in patients with persistent TNBC following neoadjuvant chemotherapy (NCT02348320). We report here on the safety, immunogenicity, and potential clinical impact of the neoantigen DNA vaccine strategy.

## MATERIALS AND METHODS

### Clinical trial

The clinical protocol was reviewed and approved by the Institutional Review Board at Washington University School of Medicine. Patients with persistent TNBC following neoadjuvant chemotherapy were eligible for participation. Patients with evidence of metastatic breast cancer or autoimmune disorders were excluded. Subjects enrolled into the protocol provided germline and tumor DNA samples, and consent for tumor/normal exome sequencing and data sharing in a controlled access database (dbGaP). After initial consent, some subjects were determined to be ineligible, or were excluded (insufficient tumor material for sequencing, patient withdrawal, and/or disease progression, Figure 1). Ultimately 18 subjects received neoantigen DNA vaccines.

**Figure 1.**
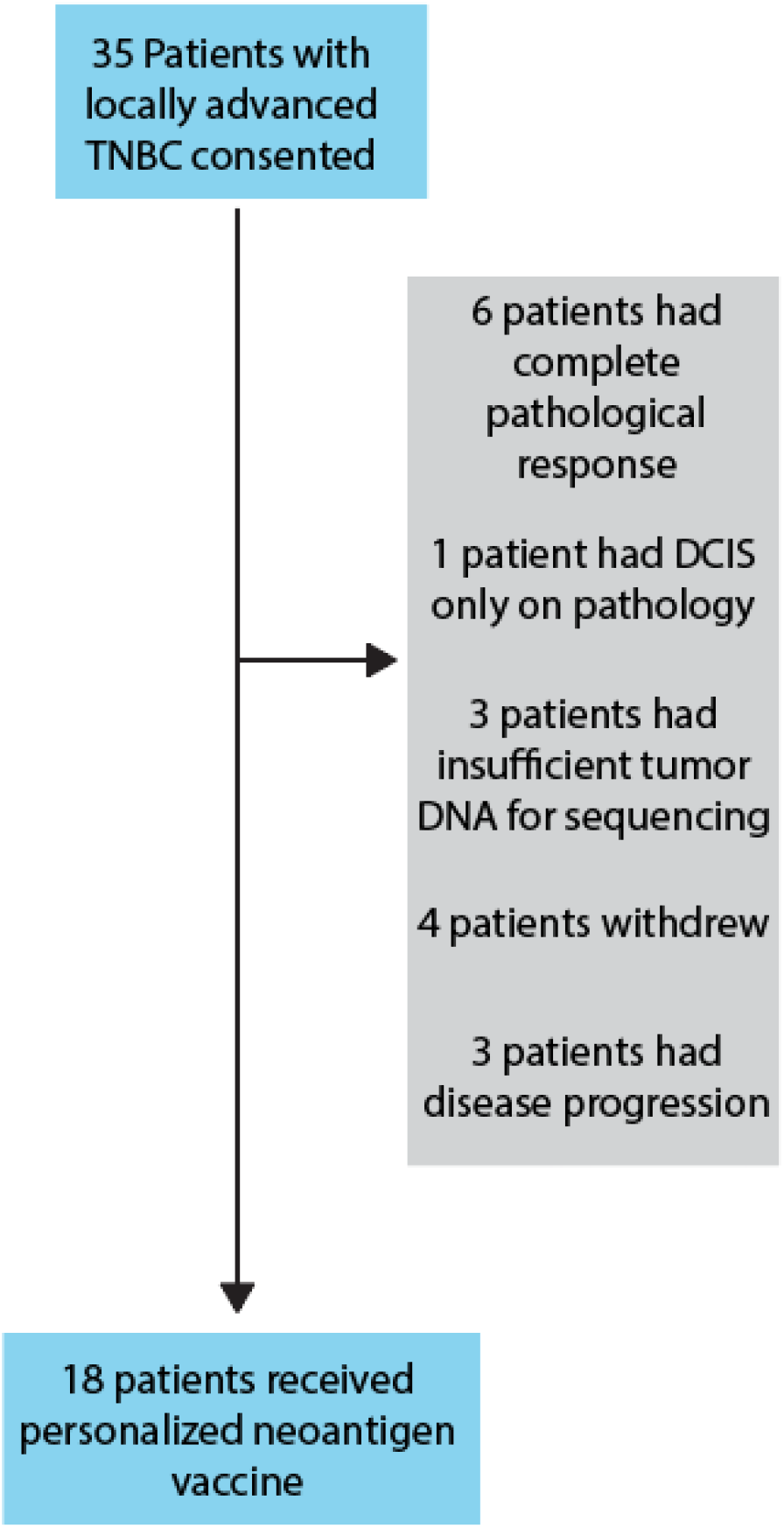
Patient selection. 35 patients with locally advanced TNBC were consented. Patients were excluded due to complete pathological response after neoadjuvant chemotherapy (NAC), insufficient tumor, patient withdrawal, and disease progression. 18 patients received personalized neoantigen DNA vaccines.

All subjects were vaccinated with 4 mg of neoantigen DNA vaccine at Day 1, Day 29 ± 7, and Day 57 ± 7. Each neoantigen DNA vaccine was administered intramuscularly using a TriGrid^TM^ electroporation device (ICHOR Medical Systems, San Diego, CA). Peripheral blood was drawn prior to each vaccination and after vaccination. Peripheral blood mononuclear cells (PBMC) were isolated through density centrifugation using Ficoll-Paque PLUS (GE Healthcare Bio-Science AB, Sweden) and cryopreserved as cell suspensions. Each subject was monitored through follow-up visits at weeks 11, 24, and 52, with additional follow-up visits or telephone contact annually thereafter. The primary objective of the clinical trial was to evaluate the safety of the neoantigen DNA vaccine strategy. Safety was closely monitored after vaccination with eight or more clinical and laboratory assessments in the first six months of the trial. Toxicity was graded according to the National Cancer Institute Common Terminology Criteria for Adverse Events (CTCAE) version 4.0. The secondary objective was to evaluate the immunogenicity of the neoantigen DNA vaccine strategy as measured by ELISPOT analysis and multi-parametric flow cytometry, both surrogates for CD8 T cell function.

### Tissue procurement and nucleic acid isolation

Archival tumor samples were obtained. H&E-stained sections were scored by a pathologist for tumor content and necrosis. Tissue blocks with over 60% tumor purity were selected, if available. DNA from PBMC was extracted using the QIAamp DNA Mini Kit (Qiagen Sciences, Maryland), and DNA and RNA were extracted from tumor tissues using the AllPrep DNA/RNA FFPE Kit (Qiagen Sciences). DNA and RNA quality were determined using an Agilent Bioanalyzer (Agilent, Santa Clara, CA), and quantitated using a Qubit Fluorometer (Life Technologies, Carlsbad, CA).

### Exome sequencing

For each subject, tumor/normal DNA samples were processed for whole exome sequencing. Libraries were prepared using KAPA Biosystems NGS kit (Roche Sequencing and Life Science, Indianapolis, IN) and captured using the IDT xGen Exome v1 panel (Integrated DNA Technologies, Inc.,Coralville, Iowa) using the manufacturer recommended procedure. Sequence data were generated as either 2 x 101 bp or 2 x 126 bp read pairs on an Illumina HiSeq instrument. DNA library preparation and sequencing were performed in a CLIA compliant space. Sequence alignment and somatic variant calling was performed as described previously [46], using an ensemble of callers and stringent filtering, followed by variant effect prediction using VEP [47].

### cDNA-capture sequencing

RNA samples were prepared using the Illumina TruSeq Stranded kit to produce cDNA, followed by cDNA capture with the IDT xGen Exome v1 panel. Both steps followed manufacturer recommended protocols with the exception of skipping the ribodepletion step on samples with low RNA yields (BRC45 and BRC10). Sequencing was performed on an Illumina HiSeq instrument, producing either 2 x 101bp or 2 x 126 bp paired-end reads. Reads were trimmed, aligned with HISAT version 2.0.5 [48] and expression was quantified using kallisto version 0.43.1 [49] and transcripts from Ensembl release 95 [50].

### HLA Typing

All patients’ HLA class I haplotypes were determined by PCR-SSOP (ProImmune, Sarasota, FL) using PBMC.

### Neoantigen Identification

We used the pVACtools [51] pipeline to identify and shortlist potential high-affinity neoantigens resulting from somatic missense mutations detected by exome sequencing. Briefly, amino acid substitutions corresponding to each of the coding missense mutations were translated into a 25-mer amino acid FASTA sequence, with up to 12 amino acids flanking the substituted amino acid on each side. For each patient, the 25-mer amino acid sequences were then evaluated through all HLA class I peptide-binding algorithms available in pVACtools to predict high affinity mutated (MT) (8-11-mer) peptides expected to bind to the patient’s HLA alleles. Matching WT sequences were also evaluated to calculate differences in binding affinities. Mutant peptides were prioritized by binding affinity (median IC50 value across multiple algorithms typically < 500 nm), sequence coverage, expression (of the gene transcript and mutant allele), variant allele fraction (preferring clonal variants to subclonal) and whether HLA anchor positions coincided with the mutation. For those candidates with a missense mutation in one of the HLA anchor positions, the fold change between mutant and wild type peptides was used to prioritize candidates with a fold change > 1. Screening was performed for incidental matches with the wild type proteome, and where appropriate peptides arising from known breast cancer driver genes were prioritized. This produced a high-confidence list of high affinity HLA class I binding neoantigen candidates for experimental validation. The list was discussed at a weekly Immunogenomics Tumor Board meeting that included physician scientists, genome scientists and immunologists. The top neoantigen candidates were selected for inclusion in each vaccine.

### Neoantigen DNA vaccine design and manufacture

Polyepitope inserts encoding prioritized neoantigens and the sequence for Ub^mut^, a mutated (G76V) ubiquitin [52] fused to the N-terminus of the polyepitope construct were synthesized by Blue Heron Biotech (Bothell, WA) and subsequently cloned into the pING vector [53]. Plasmid DNA was expressed in *E.coli* DH5α (Blue Heron) and the transformed bacteria were shipped to the Biologic Therapy Core Facility (BTCF) at Washington University School of Medicine. Bacterial cultures were expanded at the BTCF followed by lysis and DNA extraction. Each DNA vaccine was vialed at a concentration of 2 mg/mL. Before release, each neoantigen DNA vaccine underwent rigorous product release testing to assure purity, identity, and sterility. The ability to express mRNA in mammalian cells was also confirmed. The results of the product release tests were documented in a Certificate of Analysis which was reviewed and approved by both the principal investigator and BTCF staff.

### Peptides

Peptides for immune monitoring were obtained in lyophilized form at >95% purity (Peptide 2.0 Inc., Chantilly, VA). Peptides were dissolved in sterile water or in 4% DMSO dependent on the amino acid sequence. Typically, three peptides of 15 to 16 amino acids in length overlapping by 11 amino acids were pooled to encompass the ∼25 amino acid candidate neoantigen epitope encoded by the DNA vaccines.

### ELISPOT assay

2×10^5^ PBMCs were plated in each well of a 96-well round bottom plate with RPMI (with 5% human serum, 10 units/mL Penicillin-Streptomycin, 10 mM HEPES buffer, 2mM L-glutamine, 1 x non-essential amino acid). Pooled overlapping peptides corresponding to prioritized neoantigens were used to stimulate PBMCs at 25 µM, and 50 U/mL IL2 was added every 2 days. Control PBMC were stimulated with peptides corresponding to known viral antigens. On day 12, the peptide specific immune reactivity of the T cells was determined by IFN-γ ELISPOT assay as follows. Cultured T cells were stimulated with peptide-pulsed, irradiated autologous PBMC in the ELISPOT plate followed by 20 hours incubation at 37°C. Developed spots were counted in an ELISPOT reader (C.T.L., Shaker Heights, OH). Spot numbers exceeding the average number of spots observed in the absence of stimulation + 3 standard deviations of the replicates were considered significant.

### Sample preparation and DNA Sequencing for TCR

2×10^5^ PBMCs were plated per well of a 96-well round bottom plate with RPMI (with 5% human serum, 10 units/mL Penicillin-Streptomycin, 10 mM HEPES buffer, 2mM L-glutamine, 1 x non-essential amino acid). Peptides with demonstrated immunogenicity by ELISPOT assay were used to stimulate PBMCs at 25 µM followed by addition of 50 U/mL IL2 every 2 days. Control PBMC were stimulated with non-relevant peptides or media only. On day 12, cells were harvested and genomic DNA was extracted and purified from cells using the QIAGEN Blood and Tissue Kit (Qiagen, Germantown, MD). TCRβ CDR3 regions were amplified and sequenced using ImmunoSEQ (Adaptive Biotechnologies, Seattle, WA). Data were analyzed with ImmunoSEQ software and GraphPad Prism 9.

### Flow cytometry

The following anti-human monoclonal antibodies (mAb) were used for cell staining: live/dead AF488 (ThermoFisher Scientific, Waltham, MA), CD4-PerCP-Cy5.5 (clone: RPA-T4), CD8-PE (clone: HIT8a), IFN-gamma-APC (clone B27). All antibodies were obtained from BD Bioscience (San Jose, CA). Samples were analyzed on FACSCalibur Alibur (BD Biosciences, Franklin Lakes, New Jersey, U.S), and data were analyzed using FlowJo software.

### Statistical analyses

The data analysis for this study was descriptive in nature. Neoantigen-specific responses were assessed using 2-sample t-tests to compare baseline and post-vaccination PBMC samples. Progression-free survival (PFS) was defined as time from the first injection of neoantigen DNA vaccine to the date of relapse or death, whichever occurred first. Those patients alive and relapse-free were censored at date of last contact. The distribution of PFS were estimated using the Kaplan–Meier product-limit method and compared by log-rank test. All the analyses were performed using GraphPad Prism software (GraphPad, San Diego, CA, USA) or SAS 9.4 (SAS Institutes, Cary, NC).

## RESULTS

### Treatment with personalized neoantigen DNA vaccines is feasible and safe

A total of 35 patients with TNBC consented to the trial. 17 subjects were ineligible and not treated for the following reasons: complete pathologic response to standard of care neoadjuvant therapy (n=5), insufficient tumor tissue (n=4), patient preference (n=4), or disease progression (n=4). Neoantigen DNA vaccines were administered to 18 subjects (Fig. 1). Neoantigen DNA vaccines were designed and manufactured while subjects underwent adjuvant therapy (Fig. 2). After completion of adjuvant therapy, subjects received three neoantigen DNA vaccinations via electroporation at monthly intervals. In general, vaccination was well-tolerated with only one grade 3 event (hypertension) with the remaining events being grade 2 related to pain at the injection site (13 grade 2 events out of 28 total), or grade 1 mostly related to myalgia (Table S.1).

**Figure 2.**
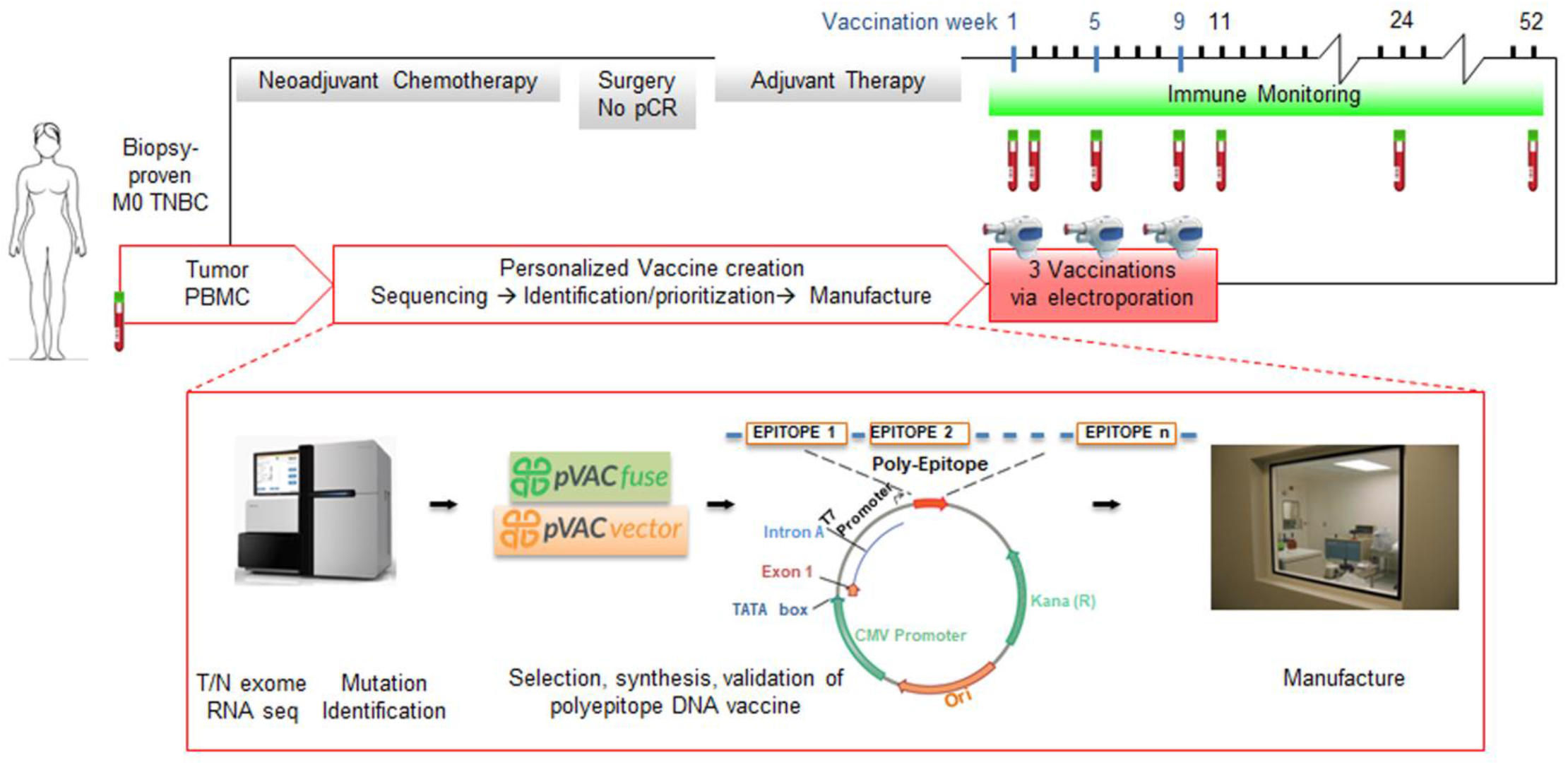
Design, manufacture and administration of neoantigen DNA vaccines for TNBC patients. Somatic mutations were identified by whole exome sequencing of tumor and germline DNA. Mutation expression was confirmed by tumor RNA-seq with cDNA capture. Candidate neoantigens were prioritized for inclusion in the vaccines on the basis of HLA binding predictions by pVAC-seq (Methods). Neoantigen DNA vaccines were administered intramuscularly using a TriGrid electroporation device.

### Neoantigen identification and vaccine design

Tumor biopsies (12 patients) or tissue obtained at the time of surgery (6 patients), and PBMC were subjected to nucleic acid isolation followed by tumor/normal exome sequencing to identify somatic mutations resulting in altered protein/amino acid sequences (Fig. 2, 3A). Tumor RNA sequencing (using cDNA capture) was performed to assess expression of somatic mutations with a median of 21.5 mutations being expressed per patient sample. Of these, a median of 8 mutations gave rise to candidate neoantigens with a predicted binding score <500nM (Fig. 3A). The median number of neoantigens ultimately included in the neoantigen DNA vaccines was 11 (range 4-20, Fig. 2, Table 1) and reflects the decision to relax binding and expression thresholds slightly to identify additional neoantigens for inclusion in some cases. 97% of the candidate neoantigens were the result of missense mutations, with the remaining neoantigens being the result of insertion/deletion or frameshift mutations (Fig. 3B). Mutations in TP53 were common, and candidate neoantigens related to TP53 mutations were present in 14/18 subjects (78%), although the location of the TP53 mutations differed among subjects (Figs. 3B, Fig. S1). Mutations in other genes that are commonly found in TNBC (such as SOX17, KMT2D, and PIK3R1) were much less frequently observed (≤17%). Of note, the data in Fig. 3B includes genetic alterations in cancer-related genes ordered by frequency of occurrence. Not all genetic alterations from such genes met the criteria for inclusion in the neoantigen DNA vaccines.

**Fig. 3.**
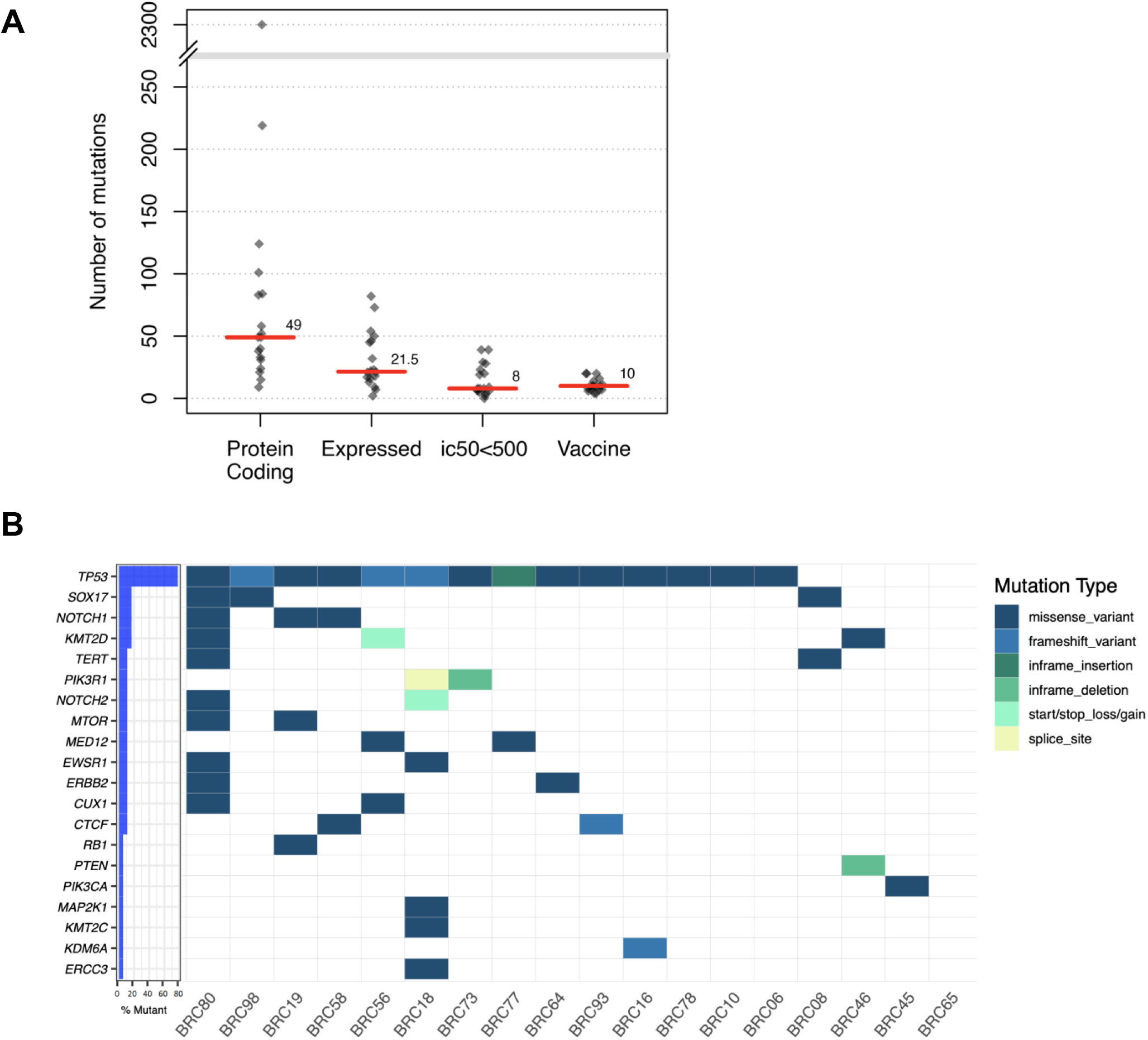
pVAC-seq analysis allows for prioritization of somatic mutations for inclusion in the neoantigen DNA vaccines. (A) Filtering plot: Counts of mutations at each stage of the mutation filtering process. Red lines and labels indicate median values. All protein-altering mutations were initially considered, followed by prioritization of those with variants expressed in the RNA, then with predicted binding affinities less than 500nm. The higher final number of mutations represented in the vaccines reflect the decision to relax binding and expression thresholds slightly to identify additional neoantigens for inclusion in some cases. (B) Waterfall plot: Mutational landscape of samples, showing protein-altering mutations in cancer-related genes. Genes are ordered by frequency and colored by alteration type.

**Table 1:**
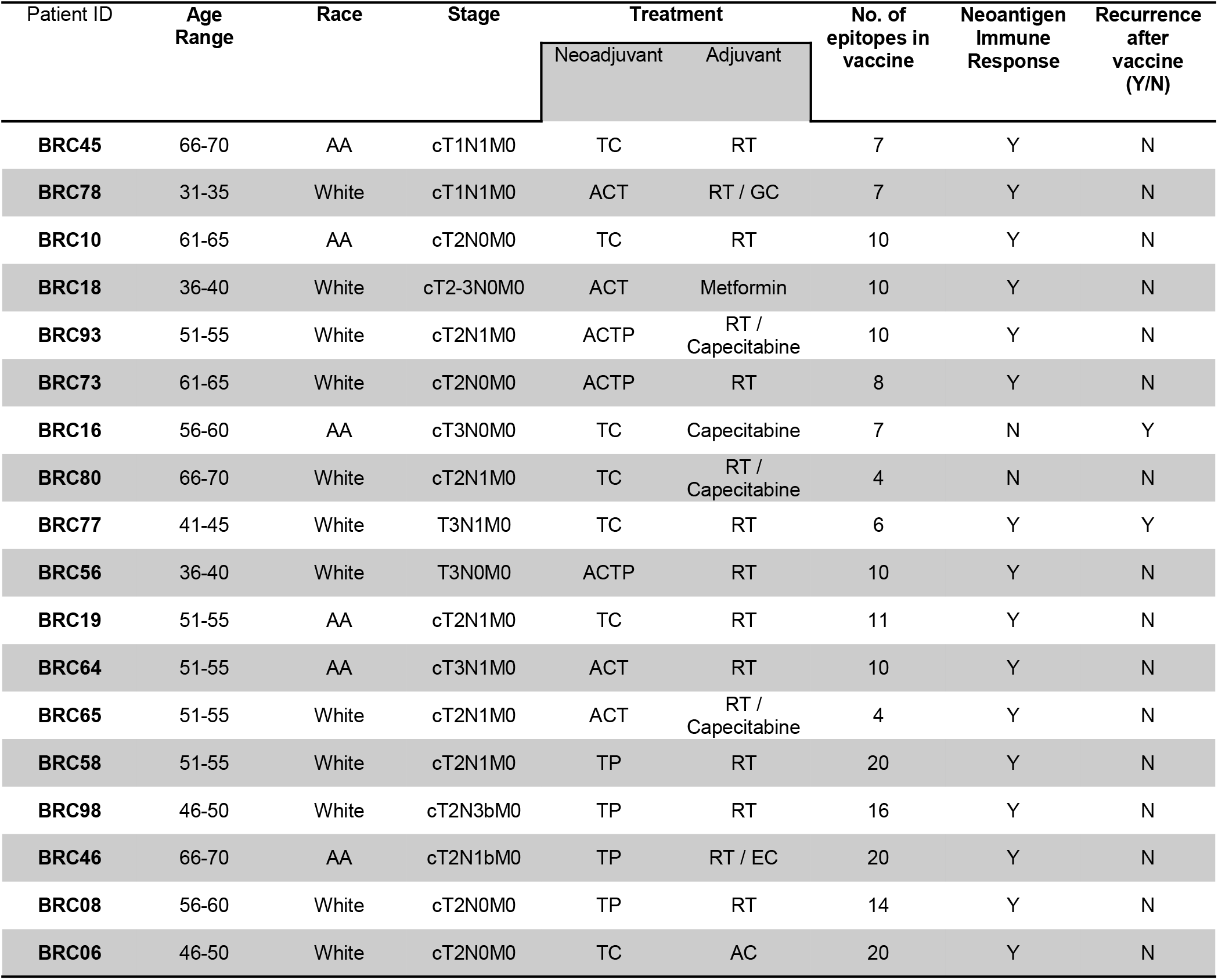
Patient baseline characteristics and immune and clinical responses to neoantigen DNA vaccination.

### Induction of neoantigen-specific responses

Immune monitoring was performed in vaccinated patients using PBMC collected at baseline and post-vaccination (Fig. 4). Immune monitoring was conducted in an unbiased manner by using overlapping peptides corresponding to each neoantigen included in the vaccine (typically three peptides 15 to 16 amino acids in length overlapping by 11 amino acids). Neoantigen-specific responses were assessed after *in vitro* culture for 12 days with the overlapping peptides (OP), followed by IFN-γ ELISPOT assay. Baseline and post-vaccination PBMC were cultured for 12 days with OP, and tested against each individual OP (Figs. 4 and S.2). A significant increase in the number of spots following vaccination was considered evidence of a vaccine-induced neoantigen-specific response. Of note, reactivity was typically the highest against the OP that included the entire predicted MHC class I epitope, whereas OP incorporating only part of the predicted epitope were poorly recognized (e.g. Fig 4A, B). To evaluate the specificity of the neoantigen-specific response, we repeated the ELISPOT assays using short peptides corresponding to the predicted mutant and wildtype MHC class I epitope. The short mutant peptides elicited equal or better reactivity than the OP, whereas the matching wild type peptide generally elicited little to no reactivity (Figs. 5 and S.2). In some cases, the response to the predicted mutant MHC class I epitope was trending lower than that against the OP (Figs. 5 and S.2). This may suggest that either the predictions failed to prioritize the most immunogenic MHC class I epitope or that the response was mediated primarily by CD4 T cells recognizing a MHC class II epitope. An analysis of intracellular IFN-γ after neoantigen stimulation is supportive of a neoantigen-specific CD4 response, showing specific responses in both CD4 and CD8 T cells (Fig. S.3). In two patients, BRC16 and BRC80, no response was detected against any of the candidate neoantigens included in the vaccine (Table 1).

**Figure 4.**
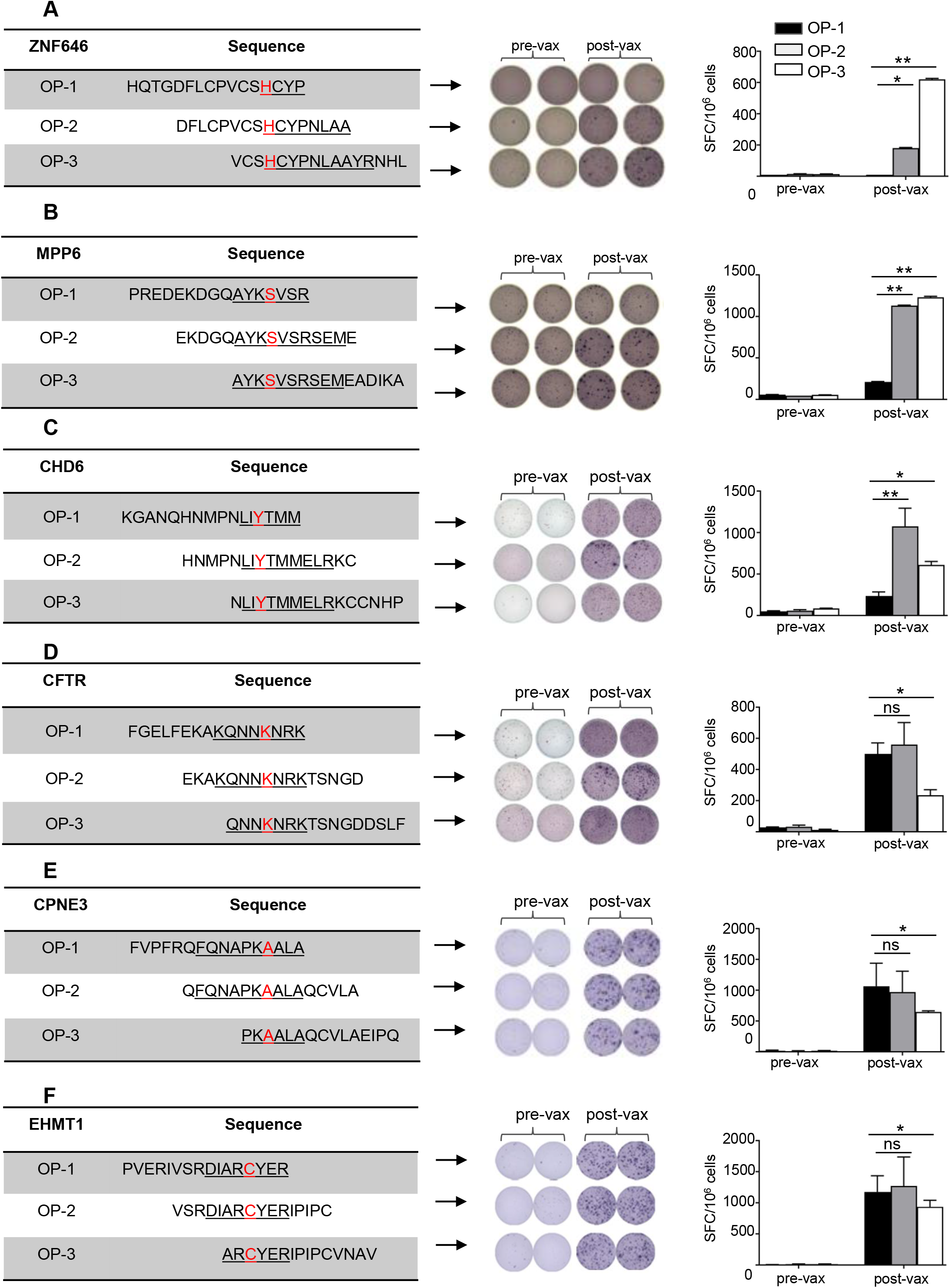
Neoantigen DNA vaccines induce neoantigen-specific immune responses. Patient-derived PBMC were stimulated with pooled candidate neoantigens for 12 days. On day 12, cells were harvested and stimulated in IFN-γ ELISPOT assays with autologous, irradiated PBMC pulsed with overlapping peptide pools of neoantigens or individual peptides. The amino acid sequence of overlapping peptides for particular candidate neoantigens is listed (left side): neoantigens for patient BCR45 in (A for ZNF646) and (B for MPP6), for patient BRC78 in (C for CHD6) and (D for CFTR), and for patient BRC18 in (E for CPNE3) and (F for EHMT1). Different overlapping peptides are indicated in color (black: OP-1; gray: OP-2; white: OP-3). The Negative controls in the ELISPOT assays included responder T cells cultured with no peptide (number of spot-forming cells per 10^6^ cells was 30–150) or irrelevant peptide (number of spot-forming cells per 10^6^ cells was 220–300). The background without peptide was subtracted from the experimental condition in each case. Data are presented as means ± SEM and are representative of three independent experiments. Samples were compared using unpaired, two-tailed Student test (*, *P* < 0.05; **, *P* < 0.01; ns, no significant difference); SFC, spot-forming cells. All T-cultures originated from pre-vaccination PBMC (pre-vax) or 2wk-post 3^rd^ vax (post-vax) PBMC; ELISPOT experiments were performed in duplicate or triplicate wells per condition.

**Figure 5.**
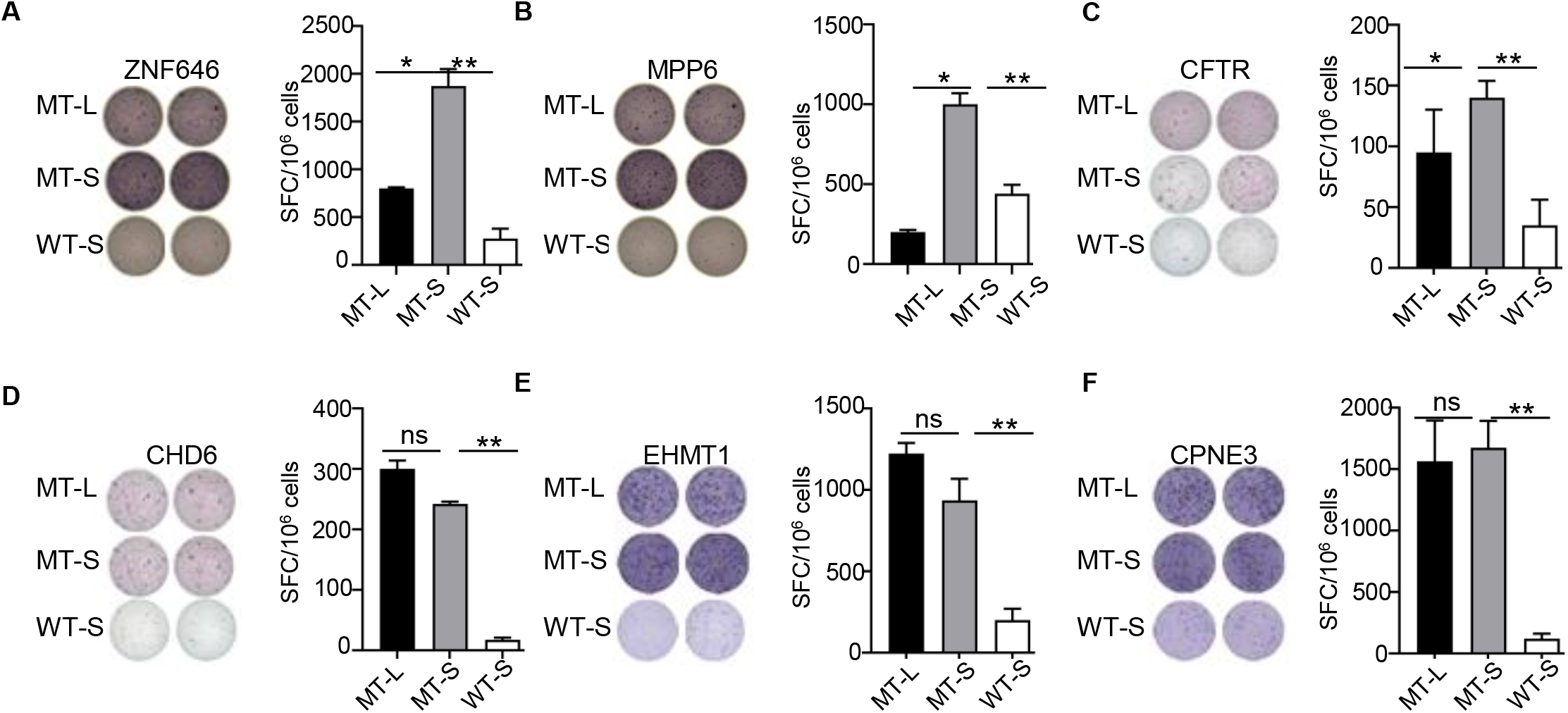
Neoantigens induced antigen-specific immune responses after in *vitro* stimulation of PBMC collected from polyepitope DNA vaccinated triple negative breast cancer patients. To confirm the specificity of the immune response induced by candidate neoantigens, T cell IFN-γ ELISPOT assays were performed on day 12 after 12-day stimulation with pooled mutant overlapping peptides by co-culturing stimulated PBMC overnight with autologous, irradiated PBMC pulsed with three pooled overlapping candidate peptides (represent one mutant gene, MT-L), individual mutant minimum (MT-S) peptide, individual wild type peptide (WT-S) and medium only or with non-related peptide as negative control. IFN-γsecretion and from representative patients BRC45 (A for ZNF646, B for MPP6), BRC78 (C for CFTR, D for CHD6), BRC18 (E for EHMT1 and F for CPNE3) against MT-L (black) peptide, MT-S (gray) peptide and WT-S (white) peptide are shown. The negative controls in the ELISPOT assays included responder T cells cultured with no peptide (number of spot-forming cells per 10^6^ cells was 30–130) or irrelevant peptide (number of spot-forming cells per 10^6^ cells was 220– 300). The background without peptide was subtracted from the experimental condition in each case. Data are presented as means ± SEM and are representative of three independent experiments. Samples were compared using unpaired, two-tailed Student test (*, *P* < 0.05; **, *P* < 0.01; ns, no significant difference); SFC, spot-forming cells. All T-cell cultures originated from 2wk-post 3rd vax PBMC; ELISPOT experiments were performed in duplicate or triplicate wells per condition.

Analysis of T cell receptor (TCR) usage before and after vaccination was performed after *in vitro* stimulation of PBMC with select neoantigens that induced a response based on ELISPOT data. Comparison demonstrated a dramatic expansion of selected TCR clonotypes after vaccination, consistent with the observed responses by ELISPOT. In some cases (EHMT1 and MUC6), multiple clones increased in frequency suggestive of an oligoclonal response, whereas in other cases (ZNF165 and CPNE3), the response appeared more monoclonal (Fig. 6). The increase in TCRβ clonotypes paralleled with intracellular IFN-*γ* production (Fig. S.3).

**Figure 6.**
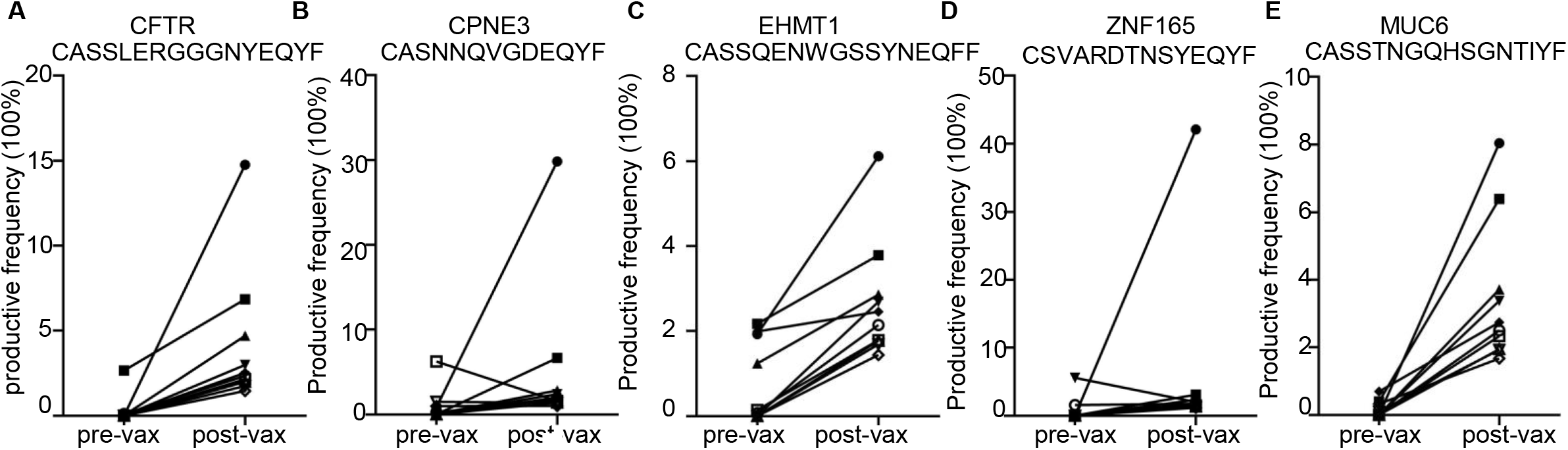
Vaccination promotes a diverse neoantigen-specific T cell repertoire. Summary of TCR-β clonotypes identified, using neoantigen-specific TCR-β CDR3 reference libraries in individual minimum stimulated PBMC obtained before and after vaccination are shown. We sequenced TCR-β CDR3 region for patient BRC78 (A), BRC18 (B and C), BRC19 (D) and BRC08 (E). Each symbol represents a unique TCR-β sequence and its frequency (%) in pre-and post-vaccine samples; P values are indicated (Wilcoxon signed-rank test).

### Preliminary evidence of clinical responses after neoantigen DNA vaccination

While not powered to formally assess clinical response, there is preliminary evidence that neoantigen vaccine treatment improved progression-free survival (PFS) compared to institutional historical controls (Fig. 7). Vaccinated patients were compared to a consecutive series of TNBC patients treated at Washington University School of Medicine between 2006 and 2010 [54]. Out of 117 patients treated with neoadjuvant chemotherapy, 60 in this series were selected as controls based on Stage II/III disease, and relapse-free status greater than 4 months after surgery. The time-0 of survival curve in the control group was reset as 4-month after surgery to account for the time required to design and manufacture neoantigen DNA vaccines). After 36 months of follow-up, PFS was 87.5 (95% CI: 72.7-100%) in vaccinated patients, compared to 49% (95% CI: 36.4-65.9%) in the institutional TNBC controls (p=0.011).

**Figure 7.**
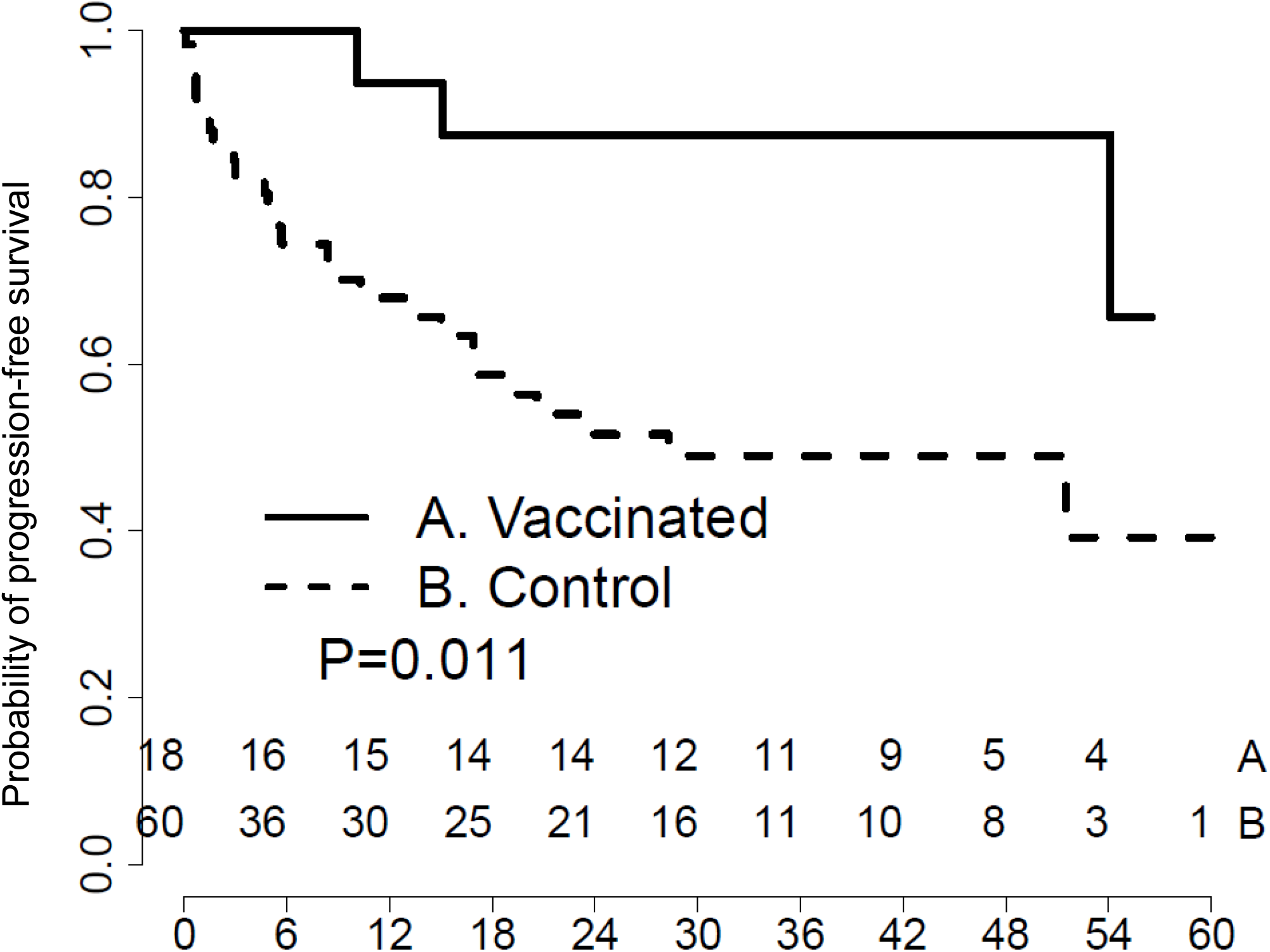
Neoantigen vaccine treatment prolongs progression-free survival compared to historical controls. Kaplan-Meier survival curve of patients vaccinated with a personalized neoantigen DNA vaccine compared to non-vaccinated historical control patients.

## DISCUSSION

Preclinical studies and early phase clinical trials have established DNA vaccines as a safe, flexible and robust vaccine platform [55, 56]. In addition to a remarkable safety profile, other advantages of the DNA vaccine platform include low cost, relative ease of manufacture compared to other vaccine platforms, and design flexibility which allows inclusion of multiple target antigens. We have recently developed and optimized a neoantigen DNA vaccine platform [5]. We demonstrated that polyepitope neoantigen constructs expressing multiple long (>20-mer) neoantigen epitopes fused with a mutant form of ubiquitin are able to induce antitumor immune responses in preclinical models. The current study is the first clinical trial that leverages this neoantigen DNA vaccine platform to target human cancer neoantigens.

The current study is also the first neoantigen vaccine study to focus exclusively on patients with TNBC, confirming the feasibility and potential of neoantigen vaccine therapy in this patient population. Prior studies have focused on high mutational burden cancers such as melanoma, non-small cell lung cancer, and bladder cancer [11, 57]. Our trial is unique in terms of targeting cancer neoantigens in a cancer with a relatively low tumor mutation burden [58]. Despite this dearth of mutations, we were able to successfully identify between 4 and 20 cancer neoantigens for each patient and produced neoantigen-specific immune responses in the majority of patients in our trial. Preliminary evidence suggests a clinical benefit as vaccinated patients had improved overall survival compared to institutional historical controls.

There is currently no effective adjuvant treatment for TNBC which is associated with a more aggressive course and has a greater likelihood of recurrence after surgery [15]. However, there is a paucity of systemic therapies available beyond chemotherapy due to TNBC’s insensitivity to hormonal therapy and/or targeted therapies. Administration of a neoantigen vaccine in the adjuvant setting is associated with several advantages. First, biopsies or surgical specimens can be used for sequencing and neoantigen identification. Second, there is a window to design and manufacture the vaccine as the patient recovers from surgery and undergoes adjuvant radiation therapy. Third, there is evidence to suggest that cancer vaccines will be most successful in the adjuvant setting, avoiding the tumor induced regulatory networks and immunosuppression that is often present with metastatic disease. Thus, neoantigen vaccines for TNBC in the adjuvant setting are not only practical, but fill an unmet clinical need.

The design of these vaccines was made possible by the pVACtools suite of computational methods for the prediction of cancer neoantigens [59]. The Tumor Neoantigen Selection Alliance (TESLA) recently compared 25 prediction algorithms and identified factors such as neoantigen MHC binding affinity, half-life, expression level, and level of foreignness as important predictors of neoantigen immunogenicity [60]. pVACtools incorporates all of these key factors identified by the TESLA consortium which likely contributed to the success rate of the vaccines in this trial.

A potential shortcoming of pVACtools as used in this trial is the emphasis on MHC class I binding given the importance of CD4 cells in reprogramming the tumor microenvironment and promoting antitumor immunity [61]. Although our neoantigen predictions prioritized binding affinity to MHC class I, we detected neoantigen-specific CD4 T cell responses in some patients. The neoantigen DNA vaccine polyepitope inserts were designed to express long peptides, 20-30 amino acids in length. Peptides of this length are preferentially processed and presented by antigen presenting cells [62], but also have the ability to bind both MHC class I and II molecules, with the ability to activate both CD8 and/or CD4 T cells. There is evidence that CD4 T cells can help CD8 T cell priming by licensing cDC1 via the CD40/CD40L interaction [63], and can help prevent CD8 T exhaustion [3, 64]. CD4 T cells also have effector roles in the tumor microenvironment including direct cytotoxicity [65], cytokine secretion, and NK cell activation.

There is also emerging evidence that many tumors have undergone MHC class I loss, but may still be susceptible to CD4 T cell-mediated immunity [3]. With recent improvements in the predictive power of MHC class II algorithms, future studiesmay be able to generate even more effective vaccines.

Our study focused on TNBC patients who had residual disease following neoadjuvant chemotherapy, a group with significantly worse survival compared to patients with complete pathologic response or to other breast cancer subtypes [66]. While not designed to evaluate clinical outcomes, there were only 2 deaths in the cohort of 18 vaccinated patients. This metric, coupled with the durable response measured by PFS is significantly better than institutional historical controls based on a study of consecutive TNBC patients seen at the WUSM between 2006 and 2010 [54]. In this institutional study, 87 patients had residual disease after neoadjuvant chemotherapy with survival greater than 4 months. Vaccinated patients had a 3-year progression-free survival of 87.5% (95% CI 72.7-100%) compared to 49% (95% CI 36.4-65.9%) in the control. This preliminary evidence of improved outcome provides strong support for further study of neoantigen DNA vaccines in TNBC patients.

Recent studies have demonstrated the importance of immune checkpoints and the tumor microenvironment in restraining antitumor immune responses. It is likely that in order to reach full therapeutic potential, cancer vaccines will need to be combined with other immune therapies such as immune checkpoint inhibition (ICI). We demonstrated in a preclinical model that anti-PD-L1 treatment is able to augment antitumor immunity mediated by DNA vaccine-induced neoantigen-specific immune responses [5], and we are currently testing neoantigen DNA vaccines in TNBC +/- durvalumab (NCT03199040). In a related study, we are investigating the combination of nab-paclitaxel, durvalumab, and tremelimumab +/- neoantigen synthetic long peptide vaccines in patients with metastatic TNBC (NCT03606967).

The primary objective of this clinical trial was to test the safety of polyepitope neoantigen DNA vaccines in patients with TNBC. We demonstrated that neoantigen DNA vaccines are safe and well-tolerated, with no significant adverse events. The neoantigen DNA vaccines were able to induce neoantigen-specific immune responses, with preliminary evidence of improved progression free survival. These results support further study of the neoantigen DNA vaccine platform in TNBC and other low mutation burden cancers.

## Data Availability

All data produced in the present study are available upon reasonable request to the authors

## ACKNOWLEDGEMENTS

We are grateful to Zhiwen He, Rebecca Neiman, Melissa Meredith, Katlyn Kraft, Sarah Larson, Leslie Nehring for coordinating the clinical trial, and to Drs. Anna Roshal, Amy Cyr, Julie A. Margenthaler, Rebecca L. Aft, Leonel Hernandez-Aya, Lindsey Peterson, Michael Naughton, Nusayba A. Bagegni, Caron E. Rigden, Timothy J. Eberlein for patient accrual and care. This project was supported by grants from Susan G. Komen for the Cure (KG111025), the Alvin J. Siteman Cancer Center (Siteman Investment Program grant 4035), the National Institute of Health (NIH) grant R01 CA240983, the National Cancer Institute (NCI) Cancer Center Support Grant P30-CA091842, the NCI grant U01 CA248235, the NCI training grant T32 CA009621, the Foundation for Barnes-Jewish Hospital (to SPG), and the Centene Corporation contract (P19-00559) for the Washington University-Centene ARCH Personalized Medicine Initiative.

---

**Figure S.1.:**
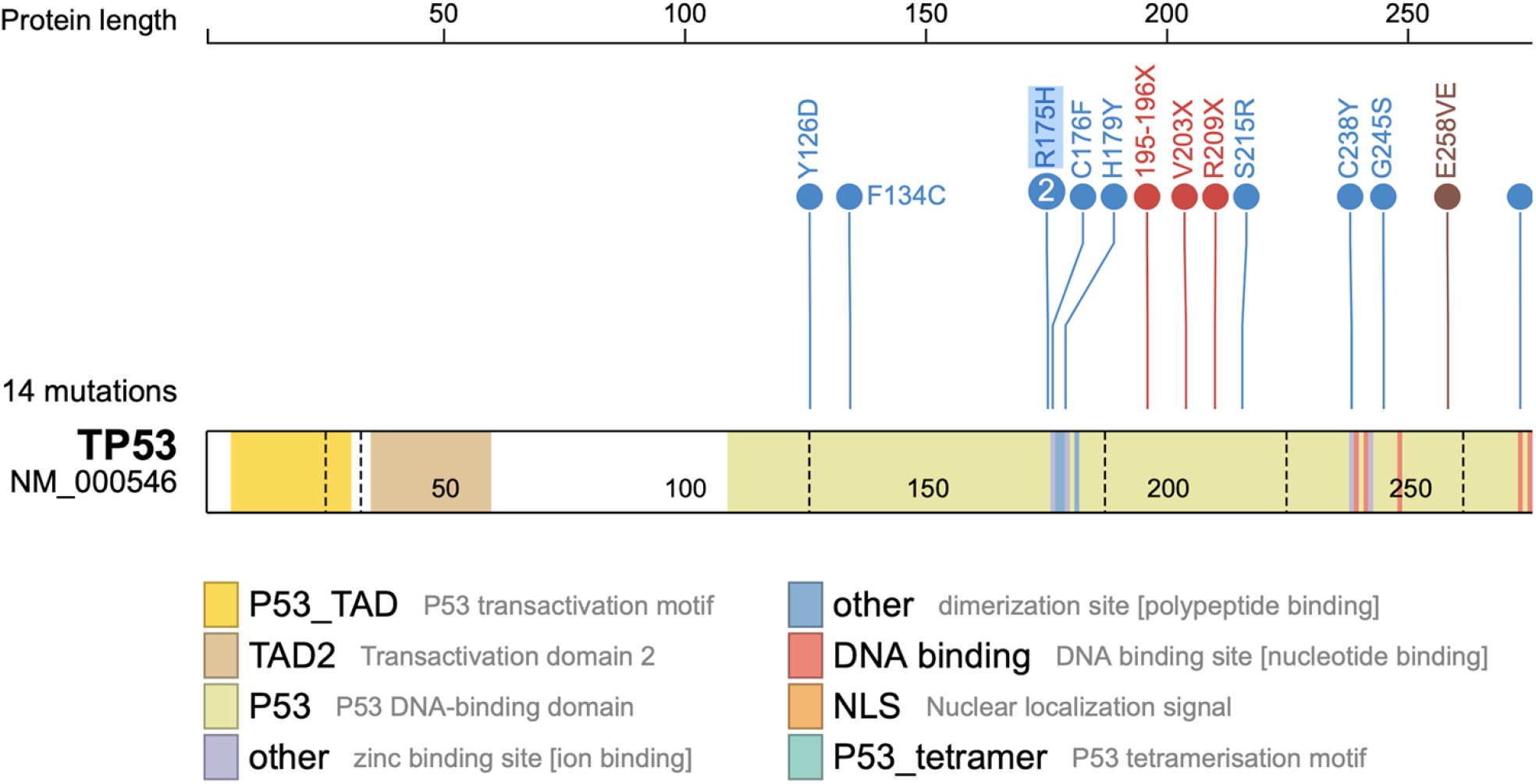
Overview of mutations occurring within the TP53 gene, with the only recurrent mutation (R175H) indicated by a “2”.

**Figure S.2.**
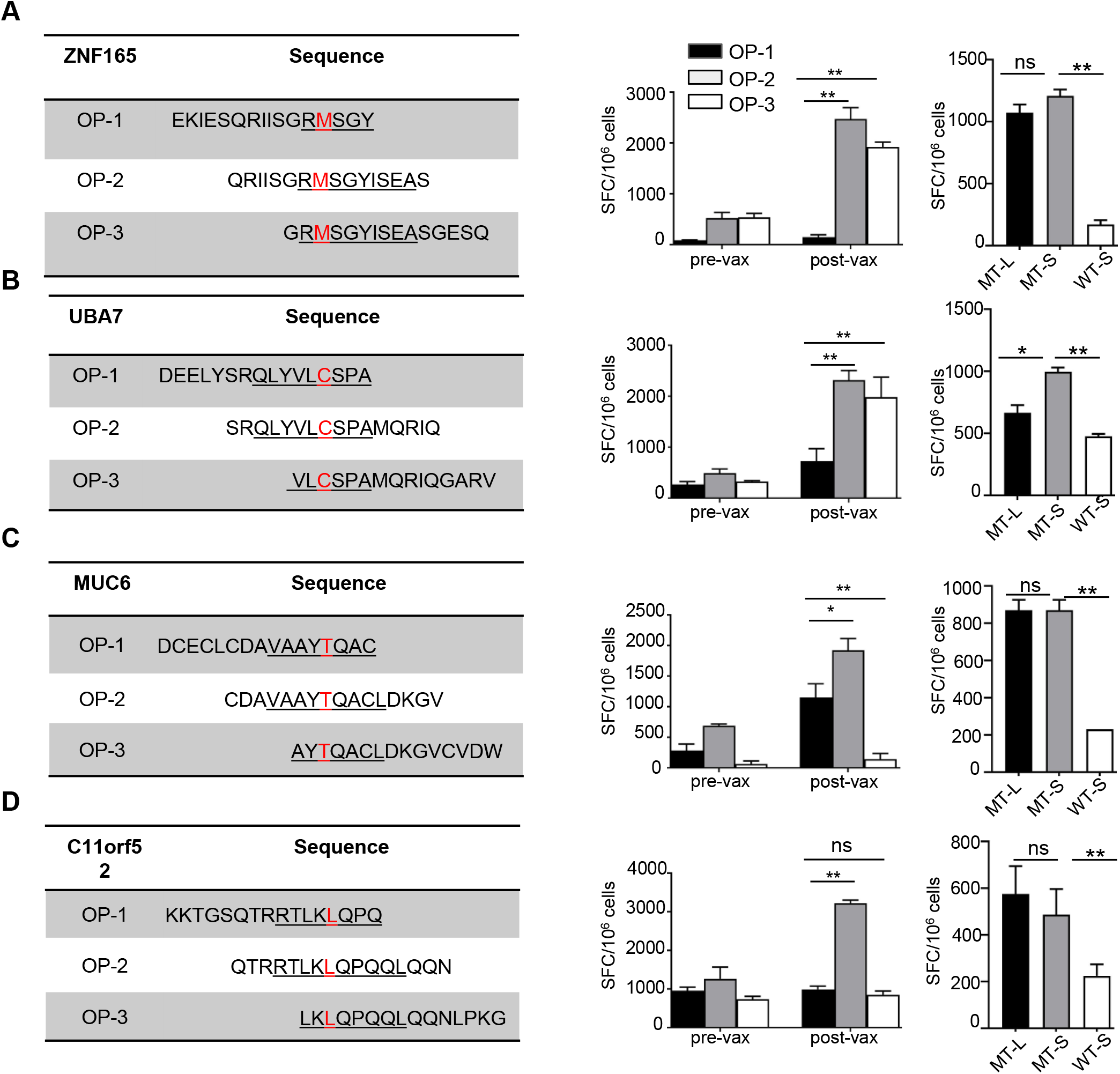
Immune reactivity to predicted candidate neoantigens before and after vaccination. PBMC at baseline (pre-vax) and after vaccination (post-vax) were stimulated with pooled OLP encoding two candidate neoantigens for 12 days. For each patient, T cell IFN-γ ELISPOT assays against pooled and individual OLP (MT-L), as well as the minimal predicted neoantigen (MT-S) and matching wild type peptide (WT-S) were performed on day 12 by co-culturing stimulated PBMC overnight with autologous, irradiated PBMC pulsed with peptide. The sequence of individual OLP from representative patients is listed with the IFN-γ secretion ELISPOT assay for patient BRC19 in (A) and (B) and for patient BRC08 in panels (C) and (D). Different OLP are indicated in color (black: OP-1; gray: OP-2; white: OP-3). The negative controls in the ELISPOT assays included responder T cells cultured with no peptide (number of spot-forming cells per 10^6^ cells was 10–120) or irrelevant peptide (number of spot-forming cells per 10^6^ cells was 120–300). The background without peptide was subtracted from the experimental condition in each case. Data are presented as means ± SEM (n =2 - 3 wells per peptide in ELISpot assay) and are representative of three independent experiments. Samples were compared using unpaired, two-tailed Student test (*, P < 0.05; **, P < 0.01; ns, no significant difference); SFC, spot-forming cells. All T-cell lines originated from 2wk-post 3rd vax PBMCs; ELISPOT experiments were performed in duplicate or triplicate wells per condition.

**Figure S3.**
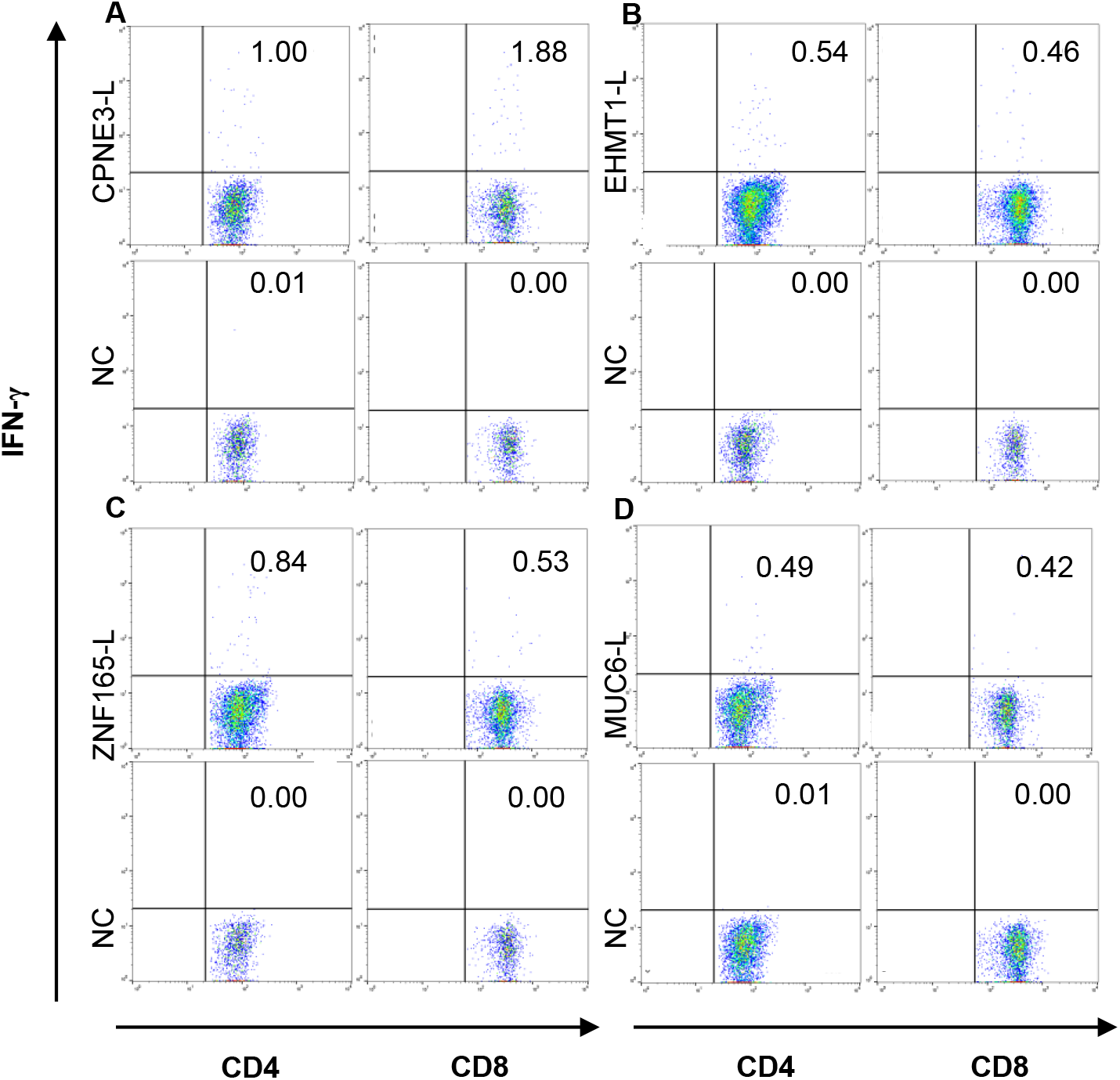
Neoantigen-specific intracellular cytokine secretion by CD4 and CD8 T cells. Post-vaccination PBMC were stimulated with pooled OLP for 12 days. For each neoantigen that induced a mutant peptide-specific immune response, intracellular IFN-γ analysis in T cells was performed on day 12 by co-culture overnight with autologous, irradiated PBMC pulsed with OLP or medium only. Flow cytometry data from patient BRC18 (A) and (B), BRC19 (C), and BRC08 (D) are shown.

**Table S.1:**
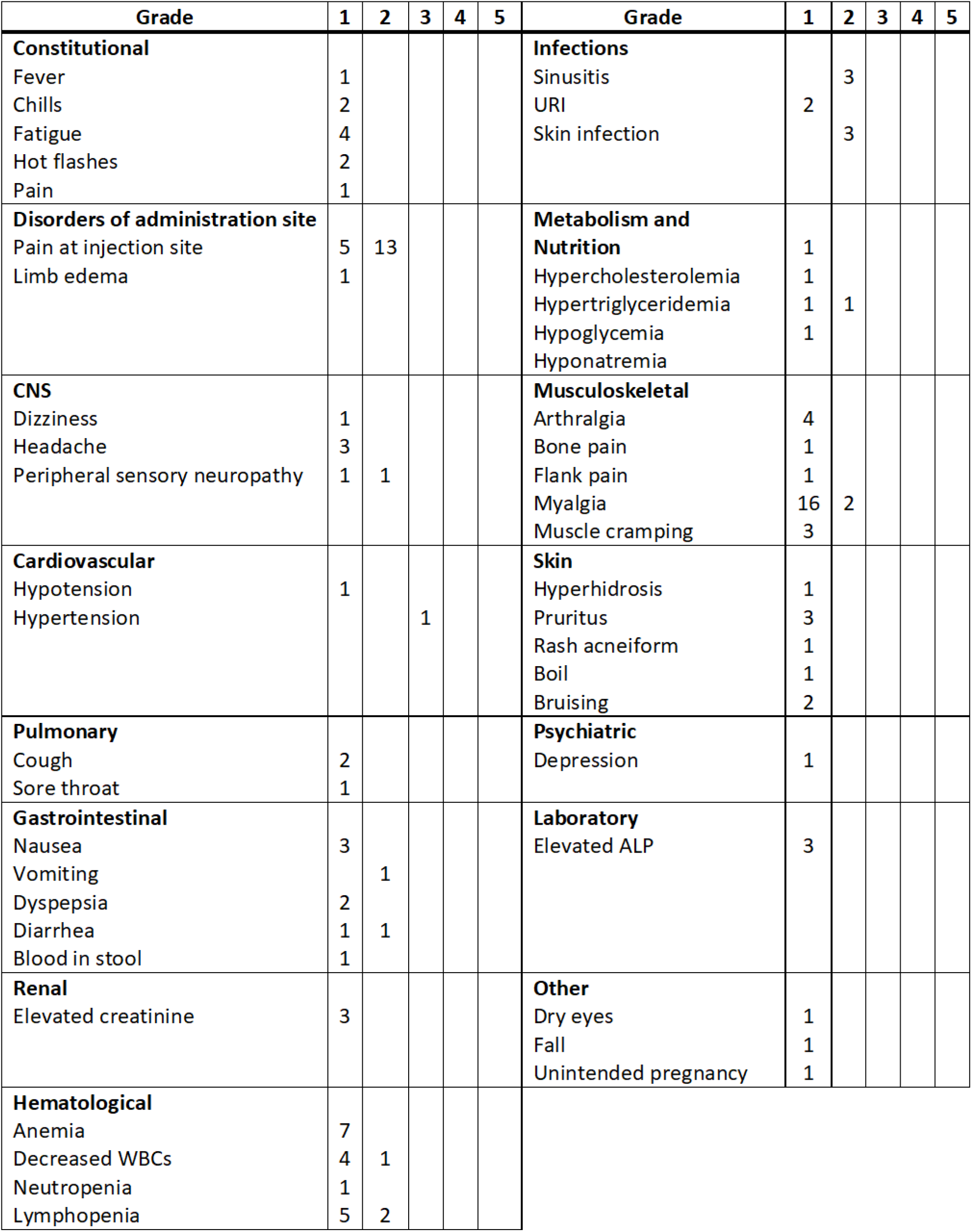
Summary of adverse events from vaccination as graded according to National Cancer Institute Common Terminology Criteria for Adverse Events version 4.0.

